# Genome-wide association study for residualized-apolipoprotein B elucidates the role of apolipoprotein B in coronary artery disease risk

**DOI:** 10.64898/2025.12.11.25341921

**Authors:** Selena K. Zhang, Satoshi Koyama, Tiffany R. Bellomo, Sara Haidermota, Whitney Hornsby, Michael C. Honigberg, Patrick Ellinor, Pradeep Natarajan

## Abstract

Apolipoprotein B (apoB) has emerged as a more accurate predictor of coronary artery disease risk relative to standard lipid measurements like low-density lipoprotein cholesterol (LDL-C). Here, we sought to characterize the clinical associations and genetics underlying apoB and LDL-C discordance. We derived a residualized-apoB phenotype, defined as observed minus expected apoB for an individual given their LDL-C level, in 239,144 individuals in the UK and Mass General Brigham Biobanks. Higher residualized-apoB was independently associated with increased risk of myocardial infarction (hazard ratio = 1.30 per standard deviation change, ΔR^2^ = 10%). Genome-wide association analyses identified 137 significant loci for residualized-apoB, including 16 unique genomic loci significant only in residualized-apoB in our study and 2 loci not previously described in lipid studies. Key loci implicated genes involved in lipid metabolism and cardiovascular disease (i.e. *NPR2*, *RORA*) and were also linked to other metabolic traits, suggesting broader metabolic relevance.

---

Over recent years, there has been increasing interest in apolipoprotein B (apoB) as a marker of coronary artery disease (CAD) risk. In addition to low-density lipoprotein cholesterol (LDL-C) as a principle target of cardiovascular disease risk modulation,^1^ numerous recent studies have affirmed the additional predictive ability of apoB on CAD risk over standard lipid parameters, such as LDL-C and non-high-density lipoprotein cholesterol (non-HDL-C).^2–7^ Mendelian randomization studies have further illustrated that, in addition to having the largest genetic effect on CAD, apoB is likely a primary lipid-related determinant in CAD.^8–10^

The apoB molecule is vital in providing structural stability to various lipoproteins, including LDL and very low-density lipoprotein (VLDL). Trapping of apoB-containing particles within the walls of arteries drives atherosclerogenesis with the degree of trapping strongly correlated with apoB concentration in the blood.^11^ The values of apoB, LDL-C, and non-HDL-C in an individual are also often highly correlated given that cholesterol makes up a large part of all apoB particles.^12^ However, despite high correlation between these markers observed in most of the population, there is often significant discordance among apoB versus LDL-C and non-HDL-C levels in some individuals. It has been found that apoB levels do not fall within a narrow specified range at a given LDL-C or non-HDL-C level and instead exhibit considerable variability at the individual patient level.^13^ Elevated apoB levels out of proportion relative to LDL-C or non-HDL-C have repeatedly been associated with increased CAD risk.^3–5,7^ The mechanisms of this discrepancy remain largely unknown, and understanding its genetic contributors may uniquely provide mechanistic insights.

Here, we sought to characterize the discordance between apoB and LDL-C to obtain mechanistic insights into apoB as an emerging risk factor for CAD distinct from LDL-C. By applying genetic analyses, we observed differential signals from the standard lipid traits studied, suggesting there may be distinct underlying pathophysiological pathways in which apoB impacts the development of cardiovascular disease.

## Results

### Clinical characteristics

A total of 216,643 individuals (mean ± SD, age 56.8 ± 8 years, 46% male) were included in the UKB analyses. Approximately 17% of the study population reported taking a lipid-lowering medication. Over median [interquartile range, IQR] 13.8 [13.0-14.5] years, 2.9% (6,346) developed incident MI. The prevalence rates of other cardiovascular disease risk factors and medical comorbidities are displayed in Table 1.

**Table 1.** Baseline characteristics. Clinical characteristics and lipid profile of the cohort populations included in the UK Biobank (UKB) and the Mass General Brigham Biobank (MGBB). Prevalence of smoking history, hypertension, and type 2 diabetes mellitus were derived from ICD codes associated with each patient in each respective cohort. *11,842 patients in the MGB Biobank had body mass index data available to include in this analysis.

| Characteristic | UK Biobank | MGB Biobank |
| --- | --- | --- |
|  | Mean (SD) | Mean (SD) |
| Age, years | 56.8 (8.0) | 55.5 (16.6) |
| Body mass index, kg/m<sup>2</sup> | 27.4 (4.8) | 27.5 (7.0)* |
|  | Number of<br>individuals (% of<br>total) | Number of<br>individuals (% of<br>total) |
| Total population size | 216,643 | 22,501 |
| Male sex | 100,309 (46.3%) | 10,014 (44.5%) |
| Lipid-lowering medication<br>prescription | 37,416 (17.3%) | 7,245 (32.2%) |
| Smoking (active or past) | 99,435 (45.9%) | 4,744 (21.1%) |
| Hypertension | 17,092 (7.9%) | 1,076 (4.8%) |
| Type 2 diabetes mellitus | 3,718 (1.7%) | 236 (1.0%) |
|  | Mean (SD) | Mean (SD) |
| Apolipoprotein B (mg/dL) | 85.0 (19.8) | 83.9 (21.5) |
| Low-density lipoprotein cholesterol<br>(mg/dL) | 67.6 (16.7) | 70.8 (19.9) |
| High-density lipoprotein cholesterol<br>(mg/dL) | 51.2 (12.6) | 55.0 (16.0) |
| Triglycerides (mg/dL) | 117.4 (51.5) | 120.2 (65.6) |

We assessed the effects of sex, lipid-lowering medication, age, type 2 diabetes mellitus (T2DM), hypertension, and history of smoking on res-apoB levels in a generalized multivariable linear model. We observed increasing age, male sex, history of T2DM, and history of hypertension were associated with increased res-apoB, whereas use of a lipid-lowering medication was negatively associated with res-apoB in UKB (Figure 1A). The generalized linear model on res-apoB was also applied to the 22,501 individuals (age 55.5 ± 16.6, 45% male) in the MGBB with similar results. As in the UKB, we again note positive effective sizes for male sex, increasing age, T2DM, and hypertension on res-apoB, correlating with known clinical pathophysiology (Figure 1A). The consistency of these associations suggest we are able to successfully capture res-apoB trends through these clinical characteristics. Moreover, regression of apoB on LDL-C yielded similar changes in apoB (mg/dL) per 1 mg/dL increase in LDL-C in the UKB and MGBB (ꞵ_UKB_ = 1.14, intercept = 7.79; ꞵ_MGBB_ = 1.01, intercept = 12.45).

**Figure 1.**
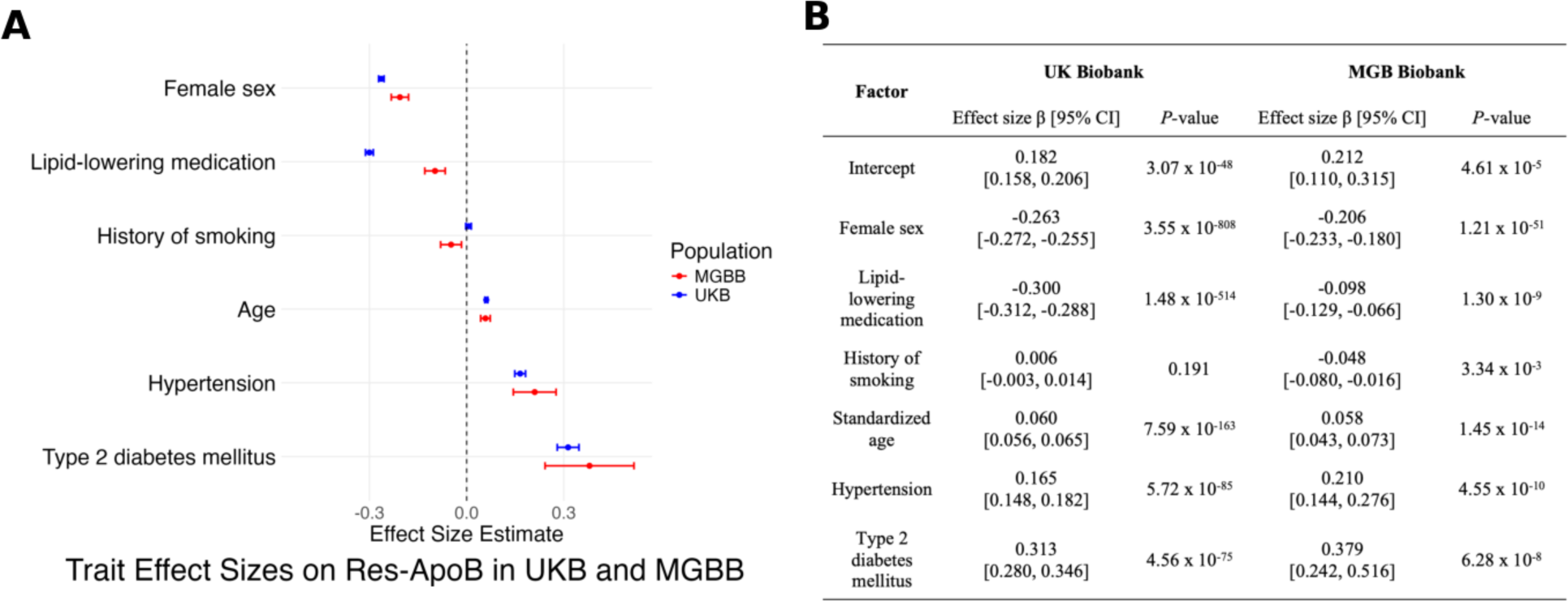
Clinical characteristics of residualized-apolipoprotein B in the UK Biobank and Mass General Brigham Biobank. A) Forest plot of effect sizes of demographic factors on residualized-apolipoprotein B (res-apoB). The effect sizes of female sex, lipid-lowering medication, age, history of type 2 diabetes mellitus, history of hypertension, and history of smoking on res-apoB in individuals from the UK Biobank (UKB) and Mass General Brigham Biobank (MGBB) are displayed in blue and red respectively. Age was standardized to mean 0 and standard deviation 1 (original standard deviations of 8.0 and 16.6 in the UKB and MGBB respectively). The effect sizes were estimated by a multivariate generalized linear model on res-apoB. B) Table summarizing effect size estimates, 95% confidence intervals, and *P*-values for each characteristic presented in A and B.

When examining its relationship to other traditional lipid traits in the UKB, res-apoB demonstrated moderate correlation with the other blood lipid traits, with a Spearman’s rho of 0.011 with LDL-C, 0.292 with apoB, -0.607 with HDL-C, 0.550 with TG, and -0.044 with Lp(a) (Supplementary Figure 2). In comparison, the correlation coefficient between apoB and LDL-C was 0.950, reflecting a highly linear relationship that has been demonstrated previously between these two lipid traits (Supplementary Figure 3). Similar patterns of correlation with res-apoB were observed in the MGBB, with a Spearman’s rho of 0.090 with LDL-C, 0.403 with apoB, -0.574 with HDL-C, and 0.533 with TG (Supplementary Figure 4). The correlation coefficient between apoB and LDL-C of 0.931 again represents a highly linear relationship (Supplementary Figure 5).

A Cox proportional hazards model was then employed to assess the impact of res-apoB on MI risk. We ran the model adjusting for different sets of covariates, including a baseline model that included age, sex, smoking history, T2DM, hypertension, Lipoprotein(a), and PCs 1-10, as well as other models that included various combinations of the traditional lipid traits (Figure 2). Addition of res-apoB to the baseline model significantly improved model discrimination for MI (ΔR^2^ = [0.0426-0.0383]/0.0426). We observed significantly increased hazard ratios (HR) for MI in the participants with higher res-apoB consistently across different models of covariates (HR range 1.044-1.301).

**Figure 2.**
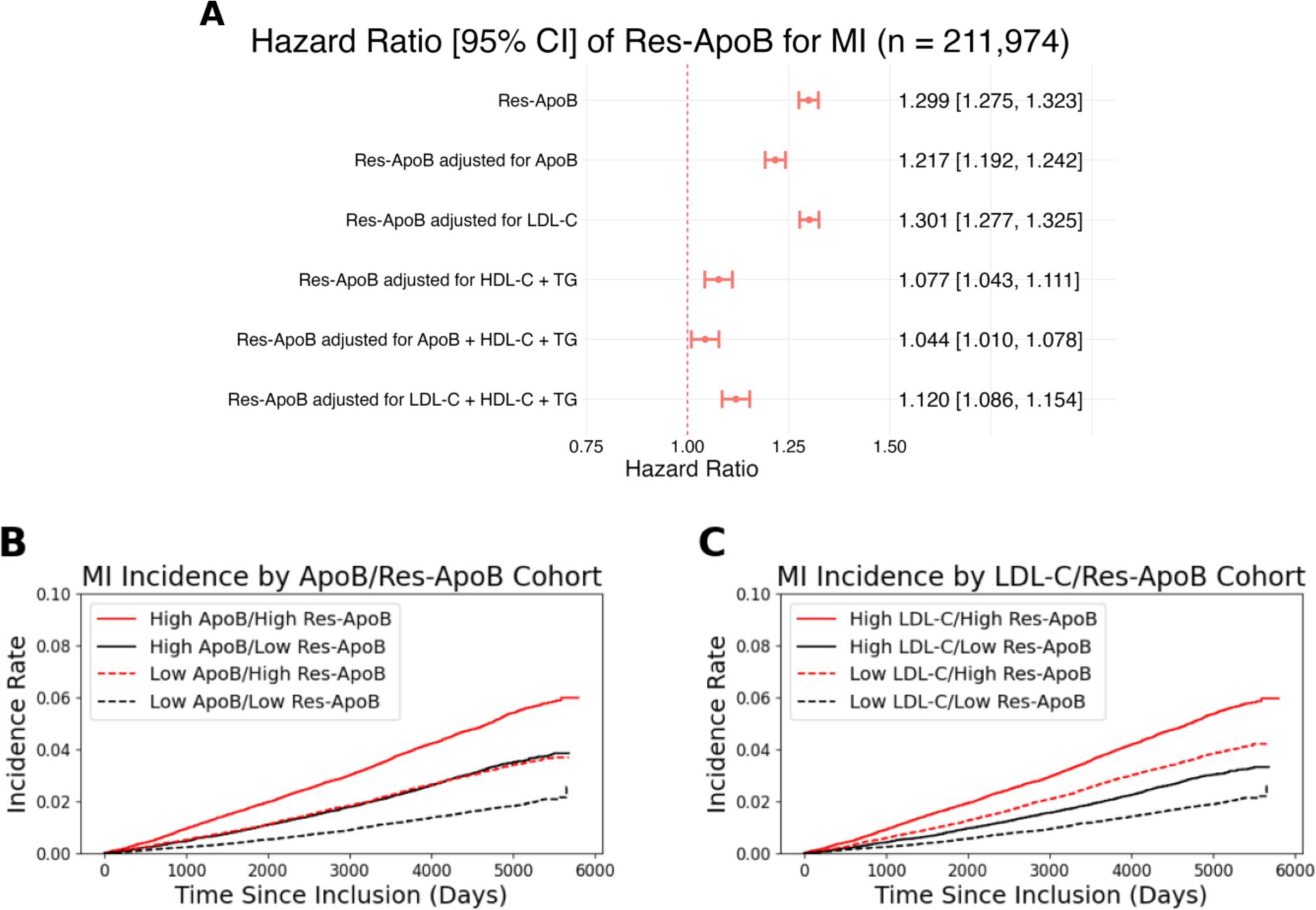
Hazard ratios and survival curves of myocardial infarction (MI) incidence. A) Forest plot of the hazard ratios of residualized apolipoprotein B (res-apoB) on myocardial infarction (MI) for several Cox proportional hazards models adjusted for different covariates. Res-apoB was standardized with mean of 0 and standard deviation of 1. The baseline model included adjusting for sex, age, the first ten genetic principal components, smoking history, type 2 diabetes mellitus, hypertension, lipoprotein(a), and res-apoB. The remaining models were adjusted for the other lipid traits as specified in addition to the baseline covariates. The points indicate hazard ratios and error bars indicate corresponding 95% confidence intervals. B) Kaplan-Meier curves displaying the incidence rate of MI stratified by apolipoprotein B (apoB) and res-apoB levels over time, excluding individuals taking a lipid-lowering medication. Cohorts were divided by median apoB level initially, before further division by median res-apoB levels. C) Kaplan-Meier curves displaying the incidence rate of MI stratified by low-density lipoprotein cholesterol (LDL-C) and res-apoB levels over time, excluding individuals taking a lipid-lowering medication. Cohorts were divided by median LDL-C level initially, before further division by median res-apoB levels.

To further understand if the increase in MI risk associated with increased res-apoB is independent of apoB and LDL-C levels, we applied Cox proportional hazards models to different stratifications of the population by apoB and LDL-C levels. The cohorts with lower-than-median apoB and lower-than-median res-apoB, or lower-than-median LDL-C and lower-than-median res-apoB were the reference groups for the two stratified analyses. Those with higher-than-median apoB or higher-than-median LDL-C, but lower-than-median res-apoB had increased MI event rates as expected (Supplementary Table 1). Relative to the cohorts with higher-than-median apoB or LDL-C, we also observed that individuals with lower-than-median apoB or lower-than-median LDL-C but higher-than-median res-apoB had elevated hazard ratios (HR_Low ApoB/High Res-ApoB_ = 1.464 [95% CI: 1.379, 1.549], HR_Low LDL-C/High Res-ApoB_ = 1.566 [1.484, 1.648]) for MI. The highest HRs were observed in the cohorts with higher-than-median apoB or higher-than-median LDL-C and higher-than-median res-apoB as expected. Incidence curves further illustrate the increased incidence of MI for cohorts driven by higher-than-median res-apoB across stratifications of apoB or LDL-C levels (Figure 2).

### Common variant GWAS in UKB and MGBB

Having demonstrated clinical relevance of res-apoB, we pursued genetic analysis to further elucidate its underlying determinants. We first separately performed GWAS of res-apoB, apoB, LDL-C, HDL-C, and TG among 216,643 European individuals from the UK Biobank and 22,501 individuals in the MGBB using NMR-based lipid measurements.^14^ The genomic inflation factor λ in the UKB was 1.282 for res-apoB, 1.297 for apoB, and 1.270 for LDL-C, suggesting an element of polygenicity and possible population stratification was present (Supplementary Table 2). LD score (LDSC) regression by the ldscr software (version 0.1.0) revealed slightly increased intercepts for res-apoB (LDSC intercept = 1.12 [SE = 0.0343]), apoB (LDSC intercept = 1.12 [0.0366]), and LDL-C (LDSC intercept = 1.09 [0.0317]).^15^ However, it is known that the LDSC intercept may be higher than 1 with increasing polygenicity.^16^ The attenuation ratios were well-controlled for all five traits, indicating the inflation was likely due to the polygenicity, and not biases introduced by miscalibration. In MGBB, the genomic inflation factor was close to 1 for all five traits (Supplementary Table 3).

The GWAS of res-apoB in the UKB identified numerous genome-wide significant variants (*P <* 5 x 10^-8^), resulting in 135 unique genomic loci in total (Supplementary Figure 6, Supplementary Table 4). We assessed the reproducibility of UKBB findings using MGBB as a replication cohort. Statistical power was computed for each of the 135 lead variants from the UKB that were present in the MGBB dataset in order to determine the appropriate set of variants for replication in the MGBB. We found that 37 of 135 lead variants had statistical power greater than 0.8 in the MGBB at an alpha level of 0.05. Of those 37, 36 exhibited concordant effect size directions, and 23 had *P* < 0.05 in MGBB (Supplementary Figure 7). The only variant with discordant effect sizes was not significant in MGBB (*P* = 0.763). These results support the consistency of the phenotyping in both biobanks and associated genetic signals observed in the UKB.

### Meta-analysis of res-apoB

We then performed a genome-wide meta-analysis in 239,144 total individuals from the UKB and MGBB to maximize statistical power in our investigation into res-apoB (Figure 2A). Significant heritability was observed for res-apoB (h^2^ = 0.114, *P* = 1.28 x 10^-35^, Figure 2B). The estimates observed were comparable to those observed for the traditional lipid traits.^17^ Consistent estimates were observed for res-apoB in the UKB (h^2^ = 0.118 [SE = 0.010], *P* = 2.94 x 10^-33^) and MGBB (h^2^ = 0.084 [SE = 0.009], *P* = 2.08 x 10^-4^, Supplementary Table 5). The genomic inflation factors were similar to those observed in the discovery analyses with λ = 1.290, LDSC intercept = 1.12, and attenuation ratio = 0.179 for res-apoB (Supplementary Table 6, Supplementary Figure 8). The attenuation ratios similarly suggest the studies were well-controlled for, with any inflation of the statistics likely due to trait polygenicity and large sample size rather than confounding.

We identified 137 unique genomic loci (*P* < 5 x 10^-8)^ with 33 lead variants and 16 loci significant only in res-apoB (Figure 2C). The 137 loci included 10 loci that were not genome-wide significant in the UKB-only discovery analysis. Of the lead variants identified in the UKB, eight were not significant in the meta-analyses due to discordant effect size directions between the UKB and MGBB (five out of eight) or only marginal significance in UKB and nonsignificance in MGBB (three out of eight). We identified a comparable number of lead variants for each of the other lipid traits as was identified in the discovery analysis as well.

The top loci for res-apoB shared genome-wide significance with other traditional lipid traits in our meta-analyses. The most significant association for res-apoB was rs328 (p.Ser474Ter, *P* = 3.38 x 10^-333^, AAF = 10.1%), a stop gained mutation in *LPL* significant also in apoB, HDL-C, and TG traits in our study. The second most significant variant was rs261291 (g.58387979T>C, *P* = 4.38 x 10^-284^, AAF = 35.5%), an intergenic variant mapped to *ALDH1A2* that achieved genome-wide significance across all lipid traits studied. This was followed by rs12448528 (g.56951643A>G, *P* = 7.38 x 10^-281^, AAF = 78.2%), a regulatory region variant mapped to *HERPUD1* and *CETP* that was significant in apoB, HDL-C, and TG traits as well. The lead variants for res-apoB additionally captured 16 missense variants, including rs11591147 in *PCSK9* (p.Arg46Leu, *P* = 3.75 x 10^-29^, AAF = 1.8%), rs116843064 in *ANGPTL4* (p.Glu40Lys, *P* = 4.82 x 10^-98^, AAF = 2.0%), and rs676210 in *APOB* (p.Pro2739Leu, *P* = 9.65 x 10^-258^, AAF = 22.0%) (Supplementary Table 7).

Two of the lead variants, rs13389219 and rs2943645, were significantly associated with T2DM in prior GWAS as well (rs13389219 *P*_Res-ApoB_ = 3.12 x 10^-16^, *P*_T2DM_ = 2.15 x 10^-11^; rs2943645 *P*_Res-ApoB_ = 1.00 x 10^-21^, *P*_T2DM_ = 4.52 x 10^-10^).^18^ Both variants had concordant effect sizes on res-apoB and T2DM (rs13389219 ꞵ_Res-ApoB_ = -0.023, ꞵ_T2DM_ = -0.066; rs2943645 ꞵ_Res-ApoB_ = 0.027, ꞵ_T2DM_ = 0.055), in concordance with the positive effect size of T2DM on res-apoB in our analyses.

Of the 16 genomic regions significant only in res-apoB in our analyses, some involved variants have previously been associated with other metabolic traits. This included missense variants, such as rs738409 (p.Ile148Met, *P* = 4.80 x 10^-21^, AAF = 22%) located on chromosome 22 in *PNPLA3* (previously associated with HDL-C, triglycerides, total cholesterol, and T2DM)^19–21^ and rs145878042 (p.Leu258Pro, *P* = 7.74 x 10^-11^, AAF = 1.1%) on chromosome 12 in *RAPGEF3* (previously associated with triglycerides, BMI, waist-hip ratio, and fasting insulin).^22–25^ However, nearly all of these previous studies were performed on larger cohorts than our sample size. Further examination of non-coding variants in the res-apoB-specific loci in RegulomeDB identified 89 signals with a ranking score of 1, indicating associations with significant eQTLs or chromatin accessibility QTLs (Supplementary Table 8). One example is rs1057208, located in an active transcription start site with hundreds of transcription factor binding signals. It also exhibits accessibility in liver tissue, with significant eQTLs demonstrating decreased expression of *PLTP*, a phospholipid transfer protein, in liver (NES= -0.27, *P* = 2.3 x 10^-6^) and other tissues. Another variant, rs7140110, has also demonstrated a potential regulatory role with active enhancer and transcription chromatin states and accessibility states observed in numerous tissues. Expression experiments have revealed significant eQTLs, including decreased expression of *GAS6*, a protein involved in cell proliferation, in liver tissue (NES = -0.48, *P* = 1.8 x 10^-13^).^26^

An example of a loci specific to res-apoB in our analyses was chr15:60,085,831-61,187,613. Its lead variant, rs71888612 (*P* = 1.76 x 10^-10^, AAF = 43.8%), is an intron variant in the *RORA* gene that exhibited a negative effect size on res-apoB (ꞵ = -0.0177 [-0.0231, -0.0123]) (Figure 3). Its effect size was significant in the UKB as well (ꞵ_UKB_ = -0.0185 [-0.0242, -0.0128], *P* = 1.94 x 10^-10^), but was not significant in the MGBB (ꞵ_MGBB_ = -0.0095 [-0.0278, 0.0088], *P* = 0.30), which could be due to the smaller MGBB sample population. However, we still observe consistency of effect size direction across the UKB, MGBB, and meta-analyses. The identified region has previously been associated with HDL-C in a study of 1.6 million individuals.^22^ Although it was not significantly associated with HDL-C in this present analysis, its association with res-apoB, despite the smaller cohort size, suggests a stronger relevance of the region to res-apoB. Additionally, two of the unique genomic regions identified were not only specific to res-apoB in our study, but also represent novel loci not reported in other previous LDL-C, HDL-C, triglyceride, and total cholesterol association studies.^27,28^ These two regions and their lead variants (chr4:177050041:TTAA:T and rs1361338) were significant only for res-apoB in our analyses. The rs1361338 variant is associated with a significant eQTL demonstrating a negative effect on *NPR2* expression in liver tissue (*P* = 3.7 x 10^-6^).^29^

**Figure 3.**
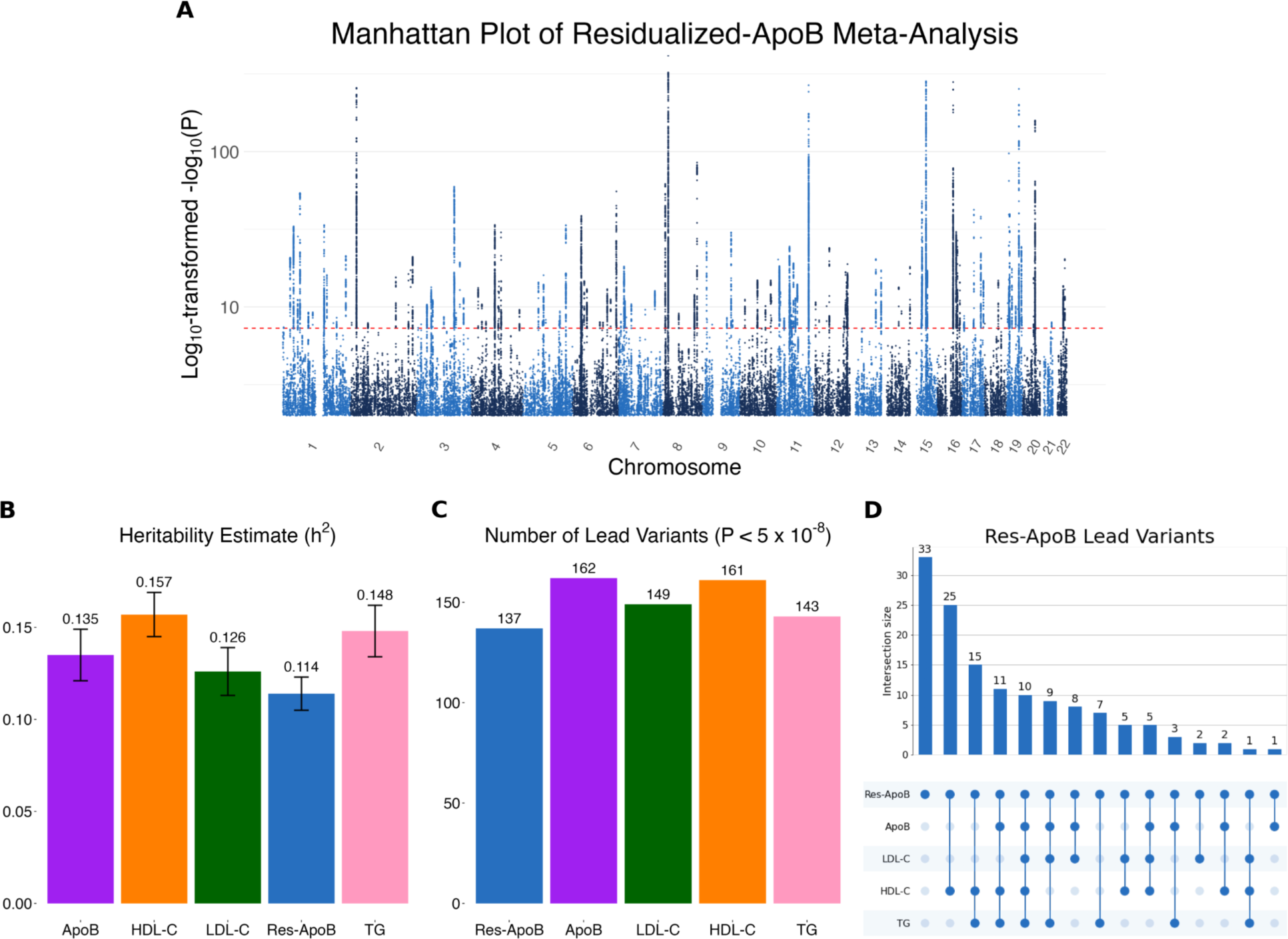
Genome-wide association study results for residualized-apolipoprotein B (res-apoB) in the meta-analyses of the UK Biobank (UKB) and Mass General Brigham Biobank (MGBB) A) Manhattan plot of genome-wide association study (GWAS) on residualized apolipoprotein B (res-apoB) on 239,144 individuals in the meta-analysis of the UK Biobank and MGB Biobank. The *P-*values are plotted as log_10_-transformed -log_10_(*P*) for visualization purposes. The dotted red line indicates the significance threshold of *P* = 5×10^-8^. B) The height of bar charts indicate the heritability estimates *h*^2^ and error bars indicate the standard error for each lipid trait in the meta-analysis. C) Bar plot of the number of lead variants (*P* < 5 × 10^-8^) identified in each GWAS for res-apoB, apolipoprotein B (apoB), low-density lipoprotein cholesterol (LDL-C), high-density lipoprotein cholesterol (HDL-C), and triglycerides (TG). D) UpsetPlot plot of the 137 lead variants identified for res-apoB with respect to whether the variants were identified as genome-wide significant in apoB, LDL-C, HDL-C, and TG. We note 33 variants significantly associated only with res-apoB and 10 variants significantly associated with all five lipid traits.

**Figure 4.**
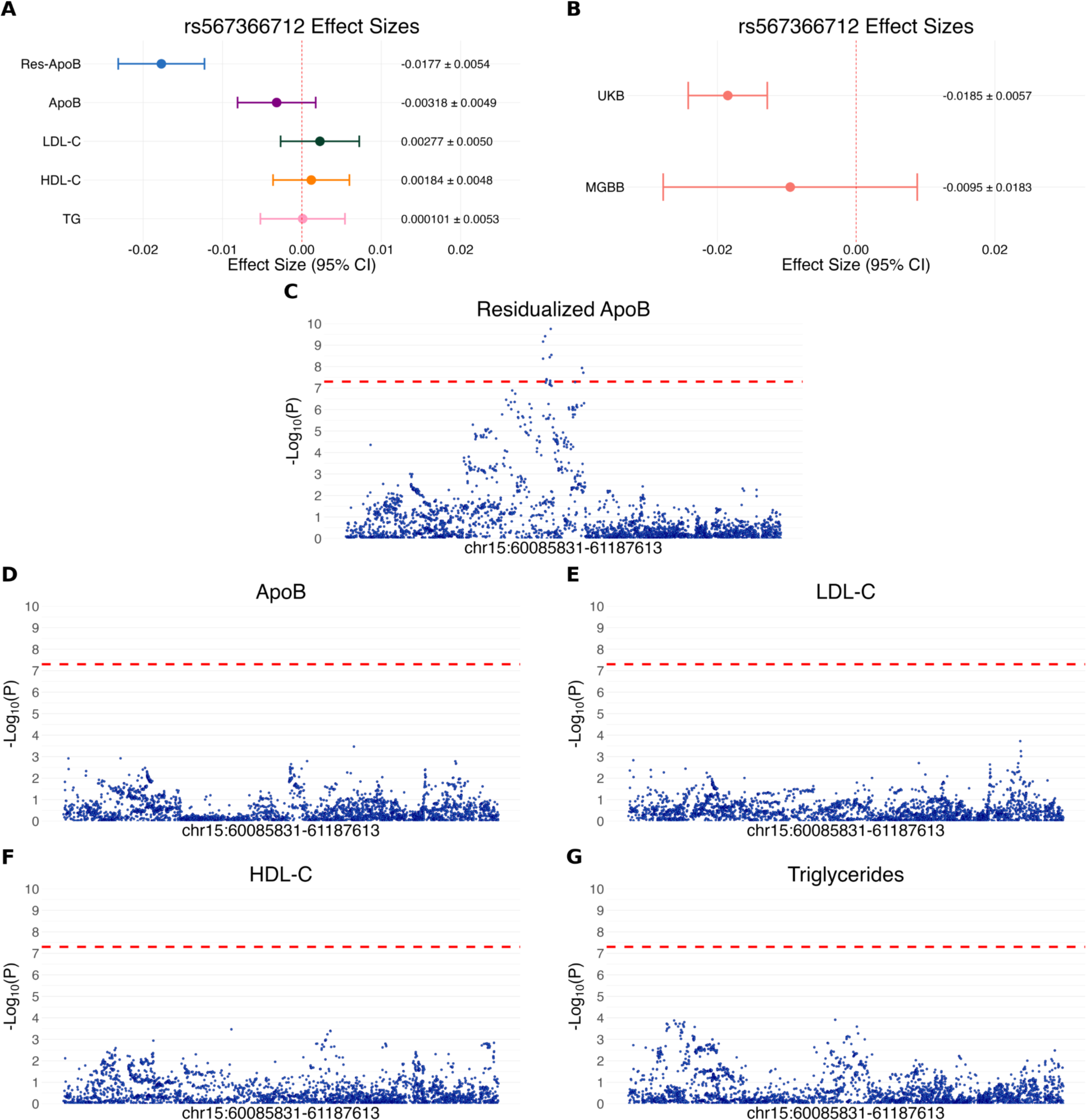
Lead variant effect sizes and Manhattan plots of *RORA* genomic region. A) Forest plot displaying the effect sizes of rs567366712, the lead variant for residualized apolipoprotein B (res-apoB) in genomic region chr15:60,085,831-61,187,613 from a genome-wide association study (GWAS) on 239,144 individuals in the meta-analysis of the UK Biobank (UKB) and MGB Biobank (MGBB). The effect sizes with 95% CI for each of the five lipid traits are shown. B) Forest plot displaying the effect sizes of rs567366712 from the UKB and MGBB GWAS. C) Manhattan plot in genomic region chr15:60,085,831-61,187,613 from the meta-analysis GWAS of the UKB and MGBB. The dotted red line indicates the significance threshold of *P* = 5 × 10^-8^. D-G) Manhattan plots of the same genomic region for apolipoprotein B (apoB), low-density lipoprotein cholesterol (LDL-C), high-density lipoprotein cholesterol (HDL-C), and triglycerides respectively.

Of the variants significant only in res-apoB, we identified 11 variants that were the sole genome-wide significant signal within their respective loci. This included 7 genomic loci significant only for res-apoB in our study. In comparing effect size directions for these variants in both UKB and MGBB, all were concordant, indicating these signals were statistically robust and likely did not represent spurious results. Further investigation revealed that 12 of the 33 of the variants specific for res-apoB, including four of the six most significantly associated variants, had discordant effect size directions on apoB and LDL-C (Supplementary Table 9). This included the most significant variant specific to res-apoB, rs738409 (*P* = 4.80 x 10^-21^; ꞵ_Res-ApoB_ = 0.0311, ꞵ_ApoB_ = 0.0030, ꞵ_LDL-C_ = -0.0078), located in *PNPLA3*. Similarly, another variant, rs11832734 (*P* = 1.72 x 10^-17^), had discordant effect sizes (ꞵ_Res-ApoB_ = -0.0379, ꞵ_apoB_ = -0.0063, ꞵ_LDL-C_ = 0.0050), illustrating that many genetic signals in our new phenotype are driven by discordance between apoB and LDL-C as expected.

We expected the genetics of res-apoB to be driven by differential effects of variants on apoB and LDL-C. We expect there to be three scenarios under which a variant is significantly associated with an increase in res-apoB, including variants that are associated with 1) a positive effect size on apoB but negative effect size on LDL-C (discordant group), 2) a negative effect on LDL-C that is greater in magnitude than its negative effect on apoB (negative group), or 3) a positive effect on apoB that is greater in magnitude than its positive effect on LDL-C (positive group). To ensure consistency in effect direction for this analysis, we aligned all lead variants such that the alternate allele corresponded to an increase in res-apoB, flipping the reference and alternate alleles, as well as inverting the sign of the effect sizes where necessary. Within our 137 lead variants, 26, 82, and 29 of the variants fell into these categories respectively. Excluding res-apoB, in the discordant and negative cohorts, the other lipid trait with the highest significance for most of the variants was HDL-C (17 out of 26 in the discordant group and 22 out of 29 in the negative group respectively), with all but one of these particular variants exhibiting a negative effect on HDL-C (Supplementary Figure 9). In contrast, the positive group had a wider distribution of the most significant lipid trait (excluding res-apoB) associated with the variants (33 with triglycerides, 26 with apoB, and 23 with HDL-C, among 82 total variants in the positive group). However, across all three cases we overall observed patterns of effect that were expected, including the variants’ negative effects on HDL-C and positive effects on triglycerides, which correlate with established literature on lower cardiovascular disease risk with higher HDL-C and higher disease risk with higher triglycerides.^13,30,31^

Lastly, to understand potential clinical implications of the genetic signals observed, we compared our variants to known gene targets for drug development. We found that 34 of the lead variants in the res-apoB meta-analyses map to previously-identified druggable genes.^32^ One example is 3’-UTR variant rs12916 located in *HMGCR*, the gene target of statin medications. rs12916 exhibited a positive effect size on res-apoB (ꞵ = 0.023, *P* = 9.76 x 10^-17^) in our GWAS. It has been associated with a significant eQTL demonstrating increased gene expression of *HMGCR* in skeletal muscle (NES = 0.18, *P* = 1.0 x 10^-10^).^29^ Missense variant rs676210 in the *APOB* gene also demonstrated a negative effect size in the genetic analyses (ꞵ = -0.116, *P* = 9.65 x 10^-258^) and exerts a negative effect on *APOB* gene expression in subcutaneous adipose tissue (NES = -0.21, *P* = 7.1 x 10^-8^). The missense variant rs11591147 located in *PCSK9* had a negative effect on res-apoB levels (ꞵ = -0.116, *P* = 3.75 x 10^-29^), but did not have significantly associated eQTLs for *PCKS9*. However, several other genome-wide significant variants within the locus (i.e. rs12117661 [ꞵ = -0.019, *P* = 5.77 x 10^-9^], rs4927191 [ꞵ = -0.017, *P* = 2.45 x 10^-8^]) displayed negative effects on *PCSK9* expression in several tissue types, including adipose tissue (rs12117661: NES = -0.49, *P* = 7.2 x 10^-18^; rs4927191: NES = -0.44, *P* = 1.9 x 10^-16^).

## Discussion

In this study, we introduced a novel lipid phenotype, res-apoB, defined as the difference between expected and observed apoB given LDL-C in an individual, to investigate discordance between these two highly correlated lipid markers. Using genetic data from the UKB and MGBB, we demonstrated that res-apoB was independently associated with incident myocardial infarction and exhibited modest correlations with traditional lipid traits. Our genome-wide meta analyses identified 137 loci associated with res-apoB, including two loci not previously identified in other lipid genetic studies. Several lead variants demonstrated regulatory effects in relevant tissues and belonged to druggable genes, supporting the biological and potential clinical relevance of this phenotype.

Our findings suggest that res-apoB captures meaningful clinical variation in cardiovascular disease risk beyond what is reflected by traditional lipid metrics. The independent association between res-apoB and incident MI, even after adjusting for traditional lipid markers, indicates that individuals with disproportionately high apoB relative to LDL-C may carry excess risk not accounted for by standard markers. Male sex, older age, hypertension, and T2DM, all features commonly associated with increased cardiovascular disease risk, were also associated with elevated res-apoB, further supporting its potential role in measuring underlying altered lipoprotein metabolism not captured in standard lipid measures. Prior studies have shown that discordance between apoB and LDL-C is associated with increased CAD risk^3–5,7^, and our results extend this literature by quantifying the discordance and demonstrating its potential value as a more personalized metric of cardiovascular risk.

The identification of res-apoB specific loci in our genetic analyses also supports its biological relevance in lipid and cardiometabolic pathways. Several associated variants were located in or near genes with established roles in lipoprotein metabolism and cardiovascular disease, including *LPL*, *PCKS9*, and *HMGCR*. Additionally, we observed associations in genes such as *RORA* and *NPR2*, which have been implicated in elevated MI risk and cholesterol regulation, as well as carotid remodeling and valvular disease respectively.^33–36^ Notably, several of these res-apoB specific loci in our study, such as in the *PNPLA3* and *RAPGEF3* genes, included variants previously linked to other metabolic and lipid traits, despite not reaching significance for the other lipid markers studied in our GWAS. The identification of signals for res-apoB in a comparatively smaller cohort suggests res-apoB may capture specific genetic influences that are missed when examining traditional markers alone, offering new paths through which to investigate cardiovascular disease mechanisms.

The overlap of res-apoB associated variants with druggable genes further underscores its potential relevance for therapeutic targeting in lowering cardiovascular risk. Variants in *HMGCR*, *APOB*, and *PCKS9*, which represent current targets of lipid-lowering medications (*HMGCR, PCKS9*) or have at least been shown to successfully reduce apoB and LDL-C levels (*APOB*),^37,38^ were significantly associated with res-apoB and often had expected effect size directions on both res-apoB and eQTLs in relevant tissues. These findings suggest existing pharmacologic agents may modulate res-apoB, and that future therapies designed to reduce discordant apoB levels could offer additional cardiovascular protection beyond lowering LDL-C alone.

This study should be interpreted in the context of its limitations. First, it has been previously established that the generalizability of results identified in genetic studies may be constrained due to differences in variant frequency, and consequently statistical power and linkage disequilibrium, across distinct ancestral populations.^39^ The UKB and MGBB both constitute primarily European populations, thus likely limiting our ability to theorize on the behavior and significance of res-apoB in non-European populations. Future studies should consider more diverse populations to mitigate this non-generalizability. Another limitation is the difference in patient populations between the UKB and MGBB. Our primary analysis in the UKB is skewed towards a healthier, middle-aged population, whereas the MGBB represents a sicker, older population. However, replication of signals in the MGBB supports the robustness of our findings and helps mitigate this concern. Still, the underrepresentation of younger adults in both biobanks limits our ability to evaluate the role of res-apoB earlier in life and its potential relevance for primary prevention strategies.

Our investigation into res-apoB as a novel phenotype suggests it represents a clinically and genetically meaningful lipid marker. Discordance between apoB and LDL-C is not only measurable and heritable, but also is associated with distinct genetic signals and clinical risk. As interest in apoB as an important driver in cardiovascular disease continues to mount, this study lays the groundwork for further exploration into the biological mechanisms underpinning res-apoB and into its potential utility in risk stratification in cardiovascular disease.

## Methods

### Ethics insights

The study protocols were approved under protocol numbers 2016P002395 and 2021P002228 by the Mass General Brigham Institutional Review Board. The analyses for the UK Biobank (UKB) were performed under application number 7089.

### Cohort description

We employed two large genotyped and phenotyped datasets, the UKB and the Mass General Brigham Biobank (MGBB), for the present study. The UKB is a large prospective population-based cohort in the United Kingdom of approximately 500,000 participants aged 40 to 69 at recruitment who were recruited during the years 2006-2010.^40^ In addition to providing genome-wide genetic data of the participants, the UKB also provides extensive phenotypic data, including biomarkers via study procedures, lifestyle factors, and biological measurements. The MGBB is a healthcare system-based cohort concentrated in Greater Boston, Massachusetts, consisting of patients recruited within the Mass General Brigham healthcare system. Most patients are primarily recruited at the Massachusetts General Hospital and the Brigham and Women’s Hospital. As of May 11, 2023, the MGBB had enrolled 142,238 individuals. The database includes genetic data interlinked with patients’ EHRs, which contain extensive phenotypic data.^41^

### Derivation of res-apoB

To characterize the concordance and/or discordance observed between apoB and LDL-C in different individuals, we created a new phenotype termed residualized-apolipoprotein B (res-apoB). Res-ApoB is defined as the difference between the observed and expected apoB for each individual. The expected apoB was calculated by regressing apoB against LDL-C under the assumption that apoB and LDL-C are often highly correlated (Supplemental Figure 1).^42^

### Survival analysis

A Cox proportional hazards model was used to assess the relationship between res-apoB and incident myocardial infarction (MI) cases [defined as International Statistical Classification of Diseases and Related Health Problems 10th Revision (ICD10), I21x, I22x, I23x (except for I23.7), I24x, and I25.2; International Statistical Classification of Diseases and Related Health Problems 9th Revision (ICD9), 410, 410.9, 411, 411.9, 412, 412.9; Office of Population Censuses and Surveys Classification of Interventions and Procedures, version 4; K40, K41, K42, K43, K44, and K46] in the UKB cohort. The model was adjusted for age, sex, and the first ten genetic principal components. Individuals taking a lipid-lowering medication were excluded from these analyses given these medications likely artificially lower LDL-C. Subsequent iterations of the model were also applied to partitions of the UKB cohort by apoB and LDL-C levels by separately dividing the population by the median apoB and LDL-C levels observed. Each of these cohorts was then subdivided by the median res-apoB. Age, sex, and the first ten genetic principal components were included in these models.

### Genome-wide association studies

For the initial genetic discovery analysis, the populations from the UKB and MGBB were chosen by selecting individuals with European-like genetic ancestry inferred from their genetic background.and with blood lipid measurements performed using the Nightingale nuclear magnetic resonance (NMR) biomarker platform.^14,43,44^ UKB participants were genotyped using a combination of the UK BiLEVE and UK Biobank Axiom arrays; detailed genotyping and quality-control procedures have been described elsewhere.^45^ In the MGBB, participants were genotyped using the Illumina Global Screening Array. Genotypes in both cohorts were imputed using the TOPMed reference panel and retained variants had imputation quality > 0.3. Quality control procedures for these cohorts have been described previously.^43,44^ There were a total of 38,904,953 variants in 216,643 individuals in the UKB. After removing rare variants with minor allele frequency < 0.01 (30,572,042 variants), 8,332,911 variants were used for analysis. We performed an equivalent analysis in the MGBB cohort. From a total of 14,965,047 variants, 6,055,388 rare variants were removed to yield 8,909,659 variants in 22,501 individuals included in the analysis. Genome-wide association studies were performed for res-apoB, apoB, LDL-C, high-density lipoprotein cholesterol (HDL-C), and triglycerides (TG) with a linear regression model using Regenie software (version 3.2.5) correcting for age, sex, genotyping array type/batch, use of lipid-lowering medications, and the first ten genetic principal components (PCs). Genome-wide significance was set to a threshold of *P <* 5 x 10^-8^. We then defined genome-wide significant regions by identifying every variant that exceeded the significance threshold (*P <* 5 x 10^-8^), expanding each signal by ±500 kb, and merging any overlapping windows, which produced non-overlapping intervals of at least 1 Mb around each lead variant. Finally, we represented each region by the variant with the smallest *P* value (lead variants). The summary statistics will be made available at time of publication.

### Replication analysis

We calculated the replication rate of the significant associations identified in the UKB within the MGBB. To account for the substantially smaller sample size of the MGBB, we also calculated the statistical power to detect each association at *P* = 0.05 in the MGBB data, using the effect sizes estimated in the UKB and the allele frequencies observed in the MGBB cohort. We stratified the associations by whether the power exceeded 80% when reporting replicability.

### Meta-analyses

We combined the UKB and MGBB cohorts to perform meta-analyses on res-apoB, apoB, LDL-C, HDL-C, and TG separately using Genome-Wide Association Meta-Analysis (GWAMA) software (version 2.2.2).^46^ The meta-analyses were filtered to remove variants with large heterogeneity statistic Q with *P_Heterogeneity_ <* 10^-6^, resulting in a total of 8,927,825 variants analyzed. To assess for the bias in our study including population stratification in all iterations of our analyses, we performed LDSC regression analysis using the ldscr software (version 0.1.0) and evaluated the intercept using European reference panel.^15^

### External datasets

To better understand the functional consequence underlying these variants, we applied Variant Effect Predictor (version 107) to the res-apoB-specific regions.^47^ The Genotype-Tissue Expression (GTEx) database was used to explore expression quantitative trait loci (eQTL) associated with the variants, which reports normalized effect size (NES) representing variants’ effects on gene expression in particular tissues.^29^ The consequences of non-coding variants were further examined using RegulomeDB.^48,49^ To elucidate potential clinical effects, the lead variants were assessed for their association with previously identified targetable genes, and their impact on gene expression was confirmed using the GTEx database.^32^

## Data Availability

All data produced in the present study are available upon reasonable request to the authors.

